# Factors Associated With Post-craniotomy Headache After Microvascular Decompression in Hemifacial Spasm

**DOI:** 10.1101/2022.03.07.22272045

**Authors:** Jiashang Huang, Yi Li, Li Jiang, Xin Li, Yan Zhan

**Affiliations:** Department of Neurosurgery, The First Affiliated Hospital of Chongqing Medical University, Chongqing, the People’s Republic of China; Department of Rehabilitation Medicine, The First Affiliated Hospital of Chongqing Medical University, Chongqing, the People’s Republic of China

**Keywords:** Post-craniotomy headache (PCH), Microvascular Decompression (MVD), Hemifacial Spasm(HFS), Complication

## Abstract

**Objective:** Patients who undergo microvascular decompression (MVD) often experience post-craniotomy headache (PCH), while the PCH is always neglected. This study is aimed to describe the natural course and risk factors of PCH.

**Methods:** The severity and duration of PCH in 87 patients who undergo MVD were recorded. Factors related to the severity and duration of PCH were analyzed.

**Results:** Most patients (63.2%) had at least one assessment of moderate to severe PCH. Almost all patients (92%) would gradually decrease to disappear within 7 days. A small number of patients (25.5%) had PCH at the incision, and other patients had PCH inside the head. Younger age and gas in the prepontine cistern were the salient features of patients in the severe group. Younger, higher SAS, gas in the prepontine cistern area, and postoperative fever were independent risk factors that affect the duration of PCH.

**Conclusions:** PCH is the most common and self-limiting complication after MVD among patients with HFS. Young age, temperature > 38°C after MVD within 24h, and gas around the TN are associated may predict the severity and duration of PCH.

**Significance:** This is the first study to describe the severity, duration, location, and risk factors of PCH.

## 1. Introduction

Microvascular decompression (MVD) is a widely recognized surgical method for the treatment of hemifacial spasm (HFS). It has advantages of high efficiency, low incidence of surgical complications, and low recurrence rate. The current reports on MVD are mainly focused on complications such as facial paralysis, hearing loss, and cerebrospinal fluid leakage(Lee, Jee et al. 2016). However, post-craniotomy headache (PCH) is a common complication after craniotomy, with reported incidence rates of about 60%(De Benedittis, Lorenzetti et al. 1996). Headache is so familiar feature after MVD surgery that many surgeons take for granted. But patients suffer and cost much from PCH. Therefore, We aimed to describe the nature of headaches after MVD and to identify the independent risk factors for PCH after MVD surgery. We hope that our findings could better understand PCH after MVD and would discover possible clinical interventions that might minimize the risk of PCH after MVD.

## 2. Methods

### 2.1 Patients

This study involved 89 patients with primary hemifacial spasm (HFS) who underwent MVD by the same surgeon (Dr. Yan Zhan) at the First Affiliated Hospital of Chongqing Medical University between November 2019 and November 2020. All patients were routinely examined by magnetic resonance imaging (MRI)to exclude posterior fossa tumors. The surgical method has been described in detail in our previous research(Huang, Zhan et al. 2021). What needs a special explanation is that we used a <2 cm micro-Keyhole to complete the operation. This study was approved by the ethics committee of the First Affiliated Hospital of Chongqing Medical University(2020-77). All patients signed an informed consent form before the questionnaire follow-up and volunteered to participate in the study.

### 2.2 Inclusion and exclusion criteria

Inclusion criteria: 1) primary hemifacial spasm, 2) no history of headaches, 3) understand the contents of the questionnaire and complete the questionnaire independently; exclusion criteria: 1) unable to complete the follow-up after the operation, 2) considering the occurrence of cranial Internal infection, 3) MVD was performed again within 1 week.

A total of 95 cases were included in this study at the beginning, of which 8 cases were excluded. 1 patient was unable to complete the follow-up due to postoperative large-area cerebral infarction; 1 patient was treated with MVD again on the fourth day after surgery due to the poor results of the first operation; 3 patients refused to continue the follow-up after the operation; 3 patients considered a possible intracranial infection. This study finally included 87 cases for analysis.

### 2.3 Grouping criteria

For all patients, the Numerical Rating Scale (NRS) scores were evaluated once a day to assess the PCH, from day 1 to day 7 after surgery. Our study mainly focused on two aspects: the severity and duration of PCH. For PCH severity grouping, we defined any NRS⩾4 as the severe group and other patients as the mild group. For the duration grouping of PCH, we defined patients with NRS⩾4 more than 3 days as the long-term PCH group and other patients as the short-term PCH group.

### 2.4 Data collection

We collected data on pre-and postoperative variables. The preoperative variables included age, sex, BMI(body-mass index), smoking (tobacco use within 3 months before surgery), drinking (alcohol use within 3 months before surgery), hypertension (blood pressure > 140/90 mmHg), diabetes mellitus, and anxiety was assessed using the self-rating anxiety scale(SAS)(Zung 1965). The postoperative variables included pH (the pH of arterial blood gas within 1 hour after surgery), fever (body temperature > 38°C within 24 hours after surgery), emesis, white blood cell count (at the 4th and 16th hours after surgery), gas in the prepontine cistern (as shown in Fig1.), intracranial air volume (using 3D Slicer 2.11 to calculate the volume of intracranial gas based on the results of the head CT scan 4 hours after surgery). PCH was assessed using a numerical rating scale (NRS, ranging from 0 for no pain to 10 for the worst pain).

**Fig.1.**
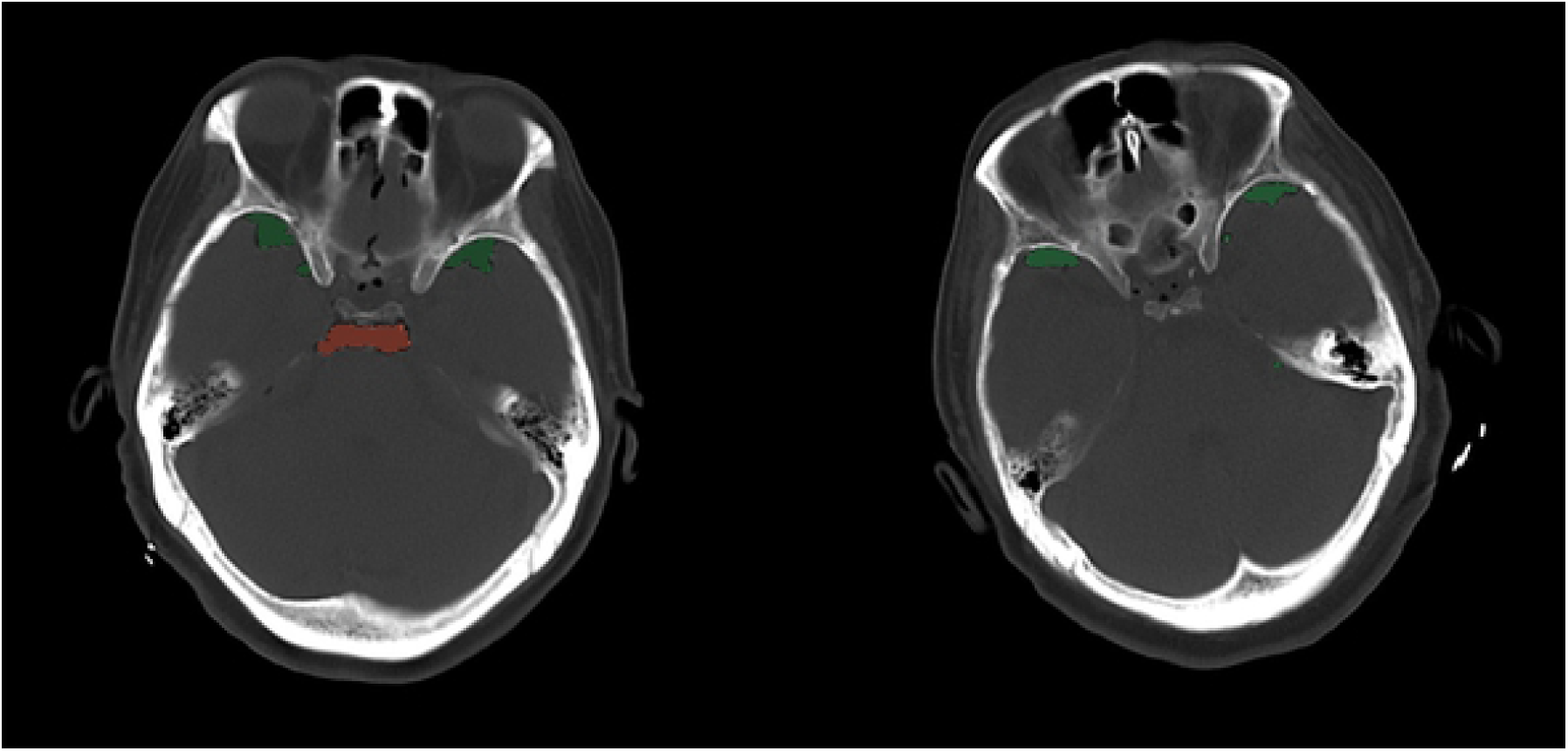
The red area shows gas accumulation in the prepontine cistern, and the green area shows gas accumulation in other parts

### 2.5 Statistical Analyses

All data were analyzed using IBM SPSS Version 24.0 (IBM Corp, Armonk, New York, USA). Categorical variables were presented as frequency (percentage). Continuous variables were presented as mean ± SD. The level of statistical significance was set at P < 0.05. Variables that were statistically significant in the univariate analysis (P < 0.05) were subjected to regression analysis.

## 3. Result

### 3.1 General information of PCH

A total of 87 patients were enrolled in this study. During the 7-day follow-up, 55 (63.2%) patients had at least one assessment of moderate to severe PCH (NRS ≥ 4 points), and the average duration of moderate to severe PCH was 1.97 days, mainly concentrated within 72 hours after surgery. The PCH of most patients(n=79, 92%) would gradually decrease to disappear within 7 days. (Fig. 2). Moderate to severe PCH was mainly distributed at the incision (n =14, 25.5%), the frontal part (n=17, 30.9%) and the temple (n=18, 32.7%), a small number of patients (n=6, 10.9%) could not identify the location of PCH (Fig. 3).

**Fig.2.**
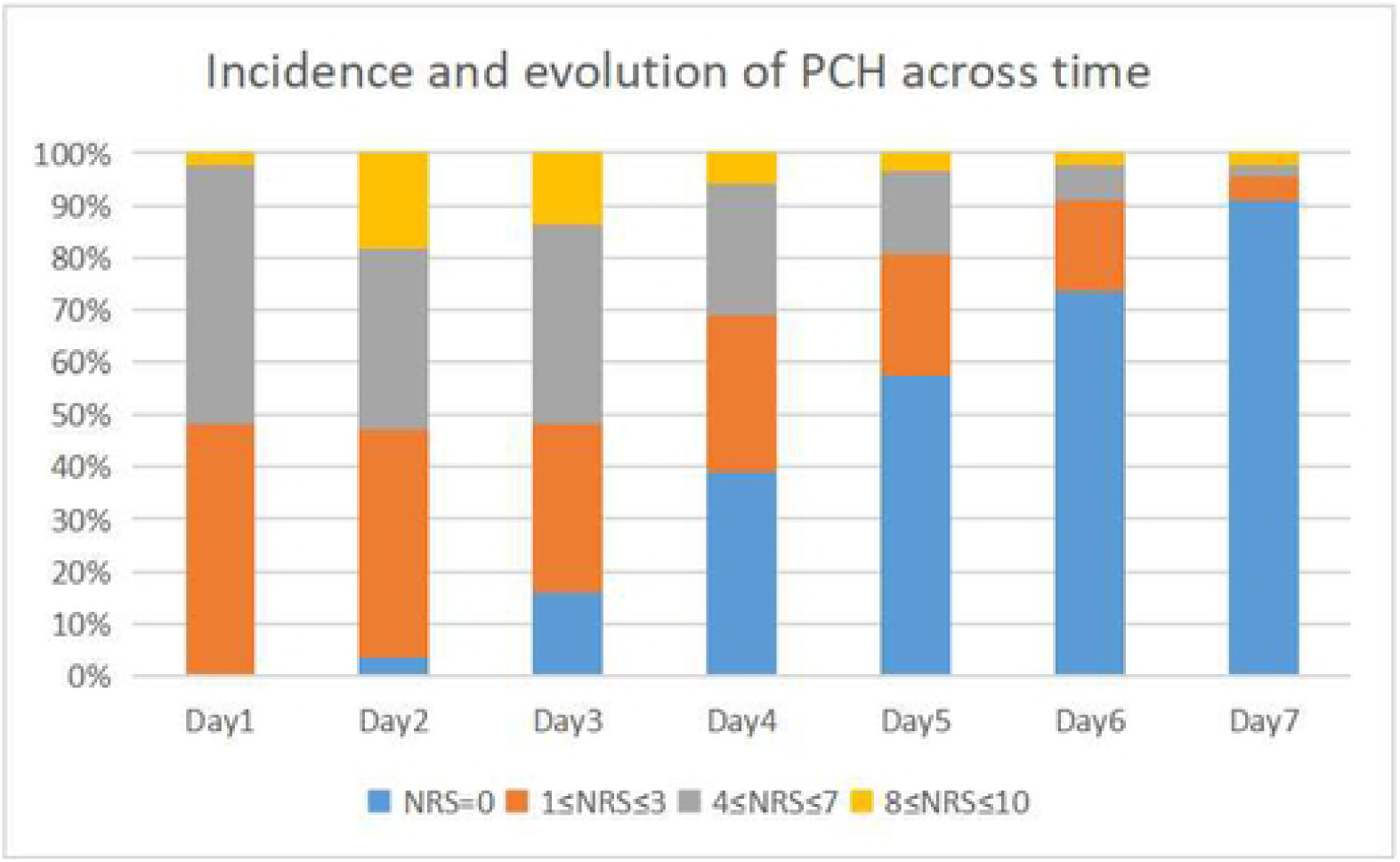
Incidence and evolution of PCH acrosstime The greatest incidence of severe pain was reported within 72 hours after MVD. The patient’s PCH gradually alleviated lo disappear within 7 days.

**Fig.3.**
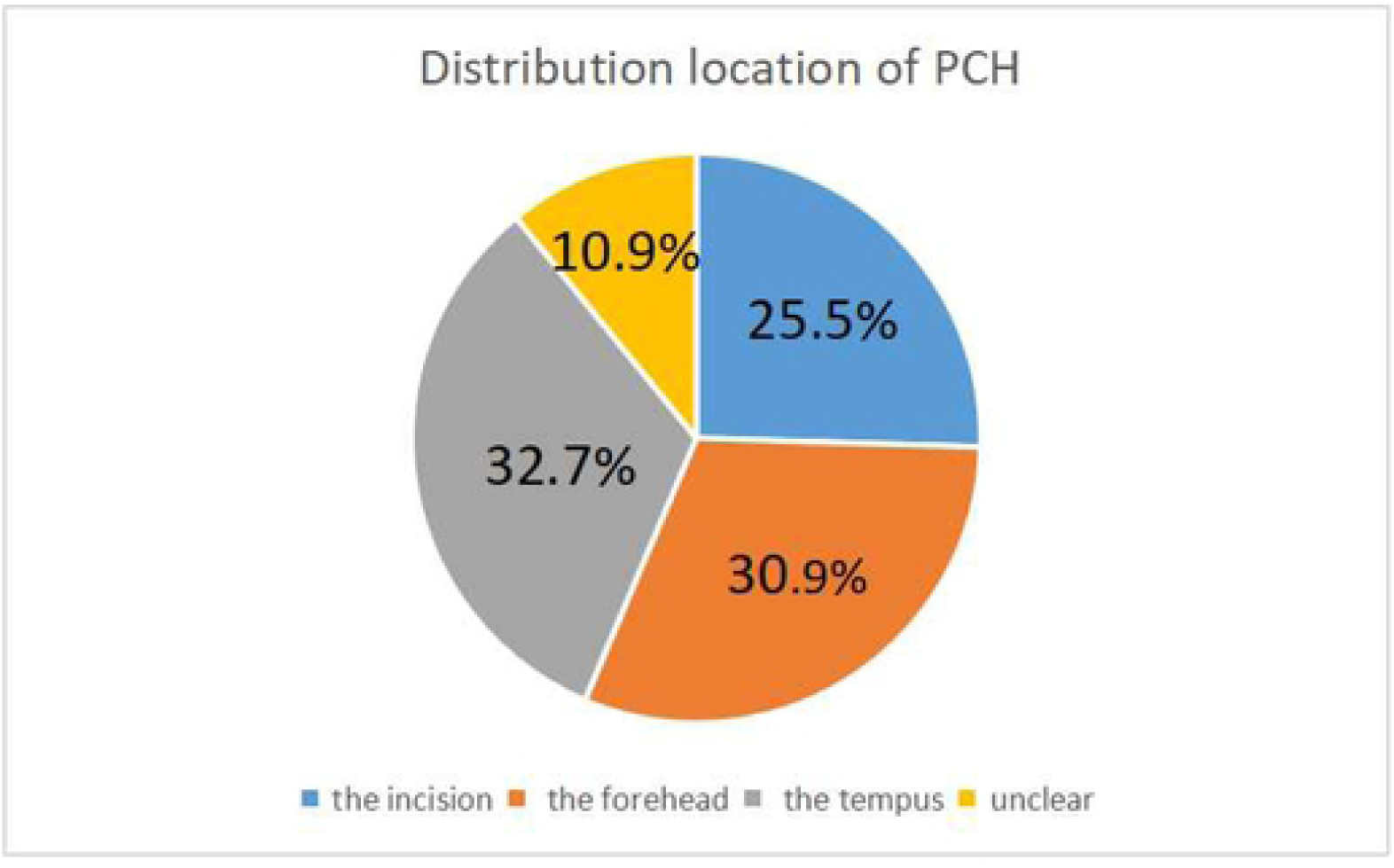
Distribution location of PCH

### 3.2 Preoperative factors

Patients in the severe group were significantly younger than the mild group (50.1±8.2VS55.8±8.3, P=0.003), while patients in the long-term PCH group were significantly younger than the short-term PCH group (47.8±8.7VS54.3±7.8, P=0.001). Additionally, patients with severe PCH had a higher score of SAS than patients with mild PCH (31.4±5.3VS28.9±5.4, P=0.040). The SAS scores of patients in the long-term PCH group were also significantly higher than those in the short-term PCH group(34.3±5.1VS28.6±4.6, P<0.001). The gender, BMI, smoking, drinking habits, diabetes mellitus, and hypertension did not significantly differ between the different PCH groups (Table 1). Regression analysis showed that age affected the severity (OR=0.930,95%CI=0.870-0.993,P=0.029) and duration(OR=0.898,95%CI=0.821-0.983,P=0.020) of PCH. In this study, the SAS score was only a risk factor for the duration of PCH (OR=1.193,95%CI=1.064-1.338, P=0.002), not a risk factor for severity of PCH (Table 3).

**Table 1.**
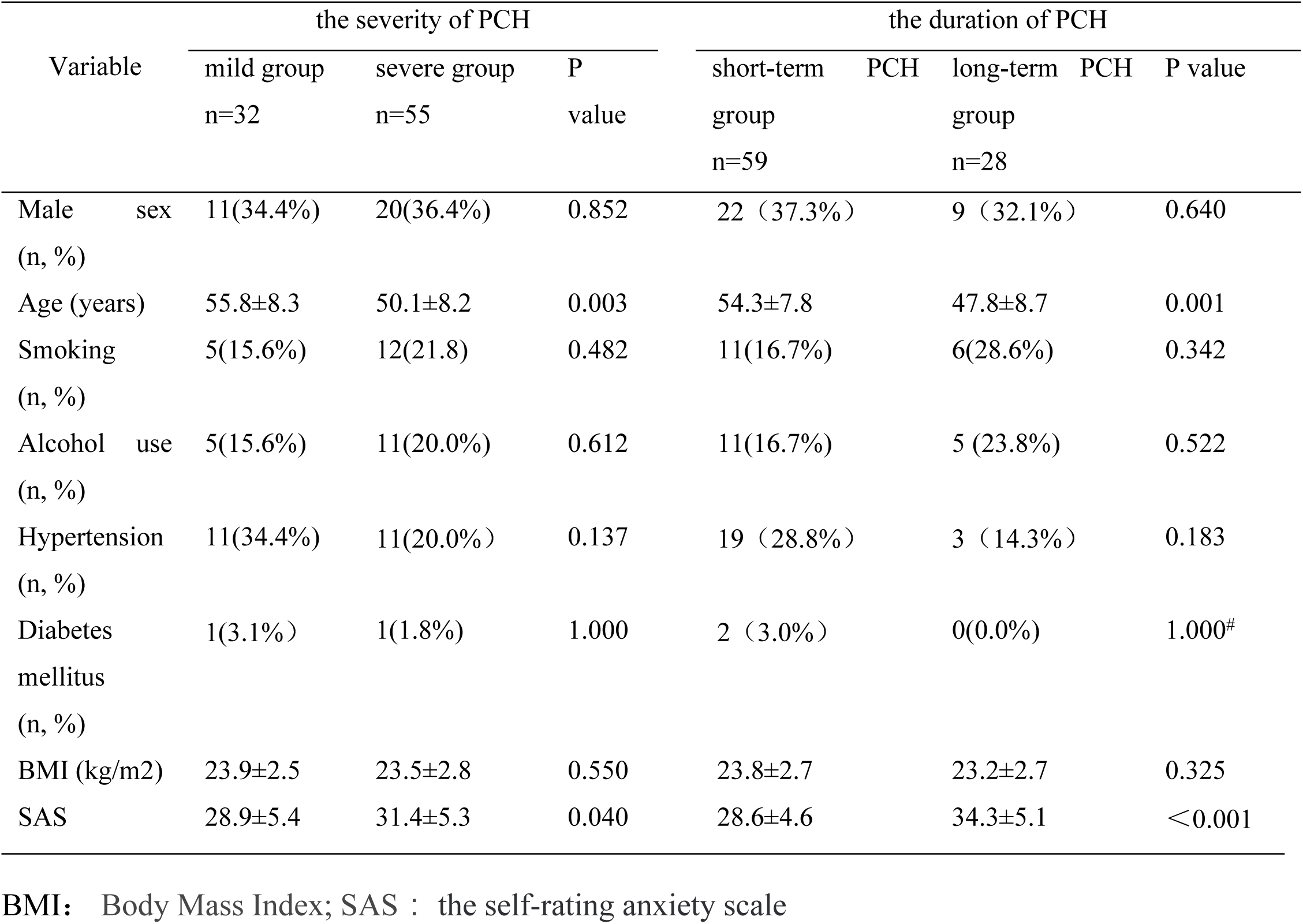
Association between preoperative factors and the severity and duration of PCH.

### 3.3 Postoperative factors

Temperature > 38°C within 24h after MVD was noted in 32.7% (18/55) of patients in the mild group and 12.5% (4/32) of patients in the severe group(P=0.036).

Meanwhile, temperature > 38°C within 24h after MVD was noted in 13.6% (8/59) of patients in the short-term PCH group and 50.0% (14/28) of patients in the long-term PCH group (OR=5.326,95%CI=1.305-21.744, P=0.020). 25% (8/32) of patients in the mild group had gas in the prepontine cistern, while 65% (36/55) of patients have gas in the prepontine cistern in the severe group (OR=3.994,95%CI=1.405-11.355, P=0.009) (Table 3). The incidence of gas in the prepontine cistern in the long-term PCH group (82.1%) was significantly higher than that in the short-term PCH group (35.6%) (OR=5.513,95%CI=1.423-12.359, P=0.013). Sodium valproate use was observed in 47.5% (25/59) of patients in the long-term PCH group and 17.9% (5/28) of patients in the short-term PCH group(P=0.025). However, sodium valproate use was similar between the mild group (28.1%) and the severe group (38.1%). No significant differences were found in any other postoperative factor between severity and duration, including pH, emesis, WBC, and intracranial air volume (Table 2).

**Table 2.**
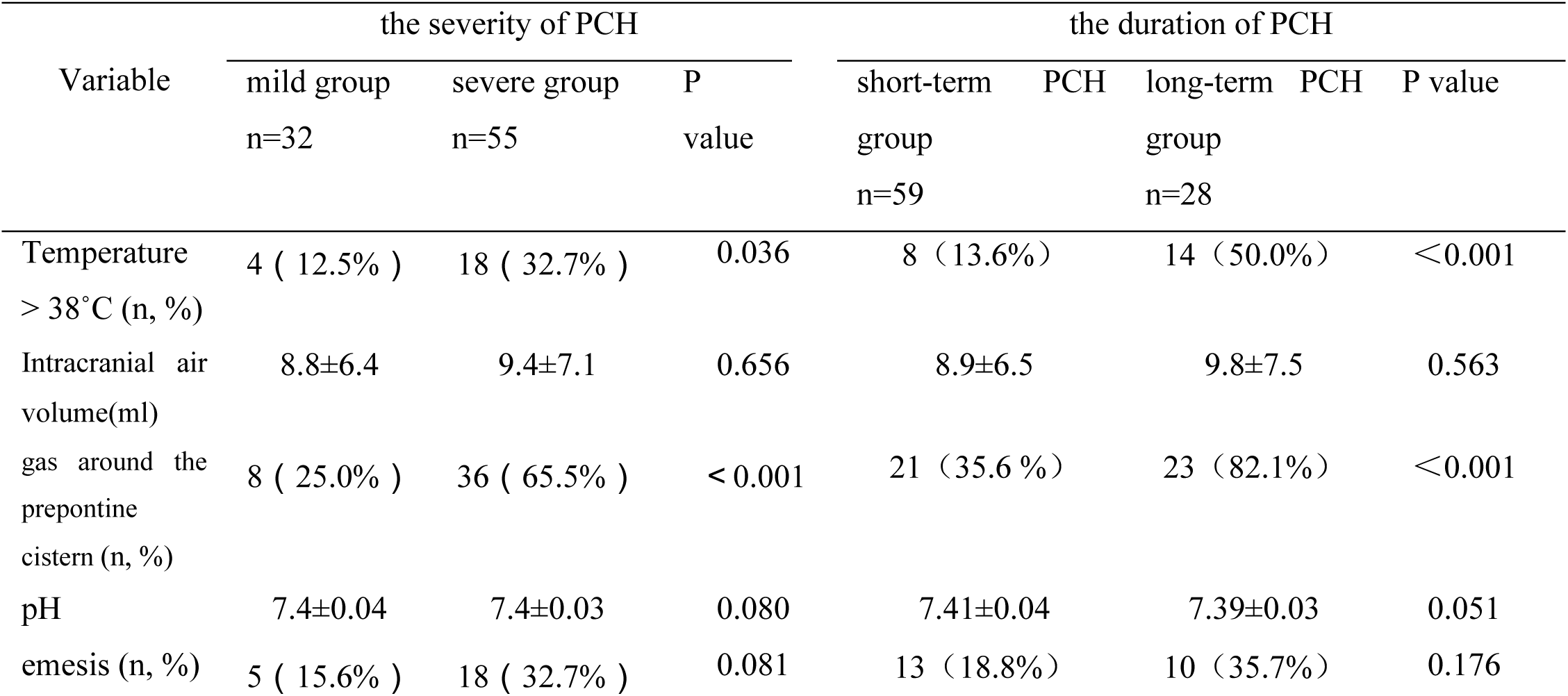
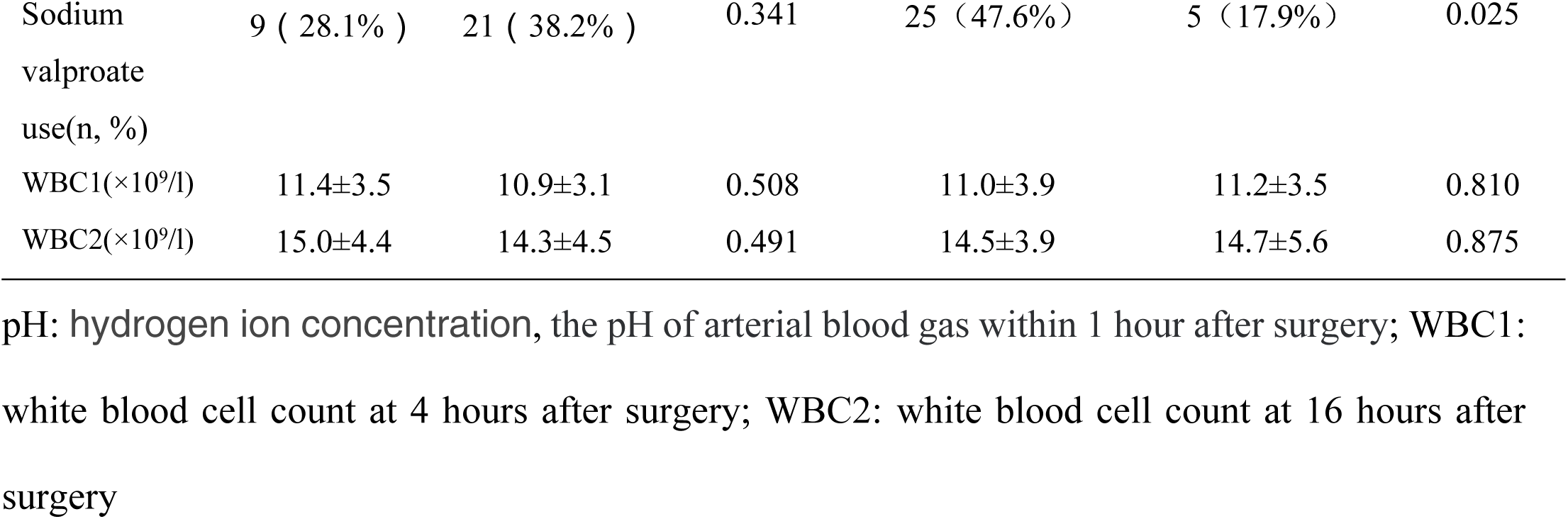
Association between postoperative factors and the severity and duration of PCH.

**Table 3.**
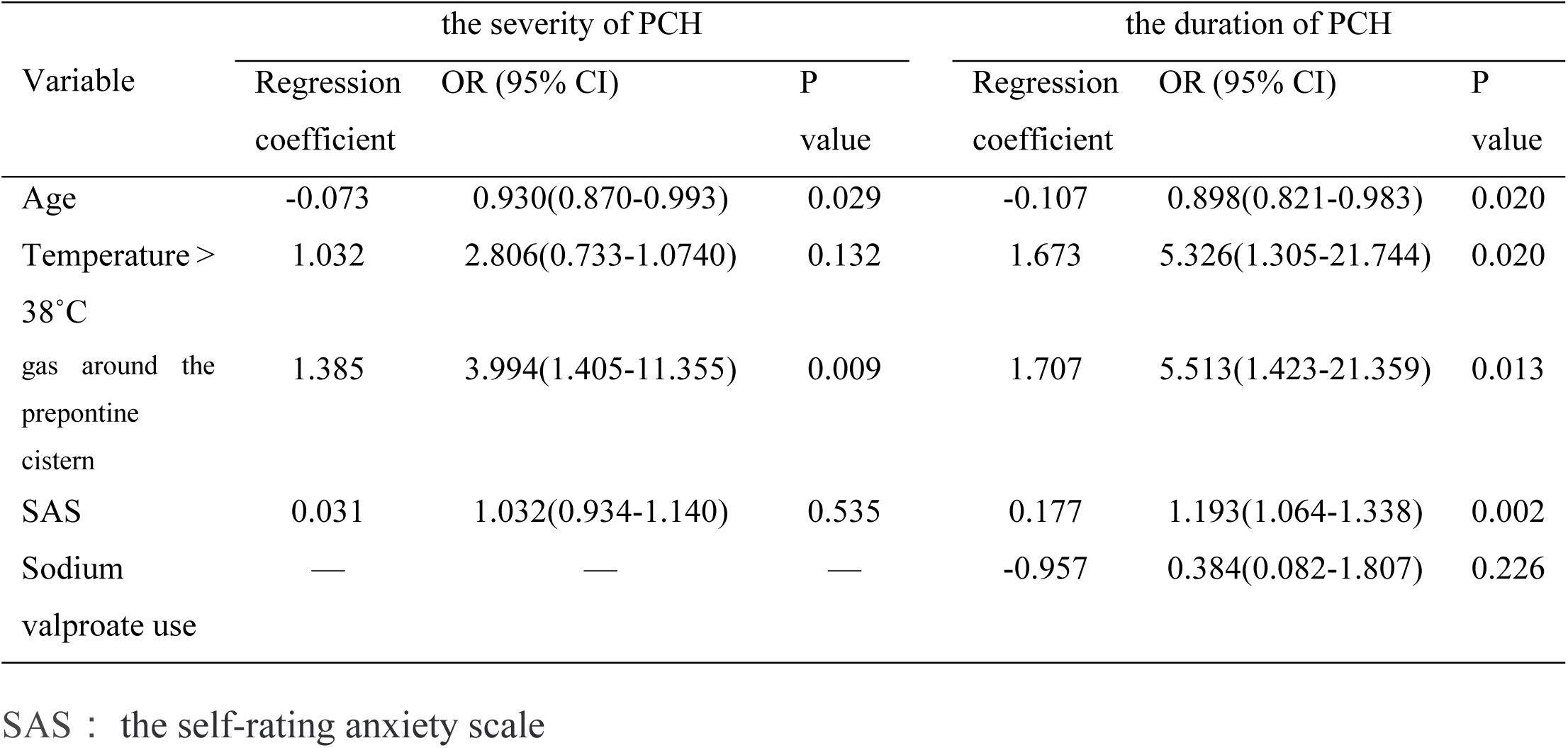
Binary Logistic regression analysis of predictors of PCH in patients undergoing Micro-keyhole MVD.

## 4 Discussion

### 4.1 Natural course of PCH

Previous studies mainly focused on PCH in patients with aneurysms or tumors but ignored PCH in patients with HFS. The purpose of our research is to record the natural course of PCH in detail and to explore independent risk factors for PCH. We look forward to increasing surgeons’ attention to PCH. In this study, 54 (62.1%) patients endured at least one time moderate to severe postoperative pain within 7 days after surgery. This means that the incidence of severe PCH is much higher than other postoperative complications of MVD(Zhao, Zhang et al. 2017). However, PCH is not listed as a postoperative complication of MVD, which indicates that PCH is ignored by clinicians. Therefore, more attention should be paid to the PCH of MVD. The latest research shows that about 60% of patients after craniotomy will suffer moderate to severe pain in the early postoperative period (within 48 hours) (Venkatraghavan, Li et al. 2016, Tsaousi, Logan et al. 2017). A prospective study(Mordhorst, Latz et al. 2010) of 256 participants who underwent elective craniotomy showed that 87% of patients reported pain within the first 24 hours after surgery, and 55% had moderate to severe pain. It can be seen that the incidence of acute headache in MVD and other types of craniotomy is similar, and the same degree of attention should be given. Our study observed that the incidence of moderate to severe PCH was 51.7%, 52.9%, and 51.7% for pain assessment at 24h, 48h, and 72h after surgery. However, the pain was reassessed 72 hours after the operation, and the incidence of moderate to severe pain was significantly reduced. Therefore, we speculate that postoperative moderate to severe headaches are mainly concentrated within 72 hours after surgery. In addition, this study showed that the number of patients experiencing moderate to severe postoperative pain gradually decreased over time, and 90.8% (n=79) of patients showed complete relief of headache on day 7, indicating that acute headache after MVD is a self-limiting headache. Therefore, we consider that giving patients appropriate analgesic measures within 72 hours after surgery can effectively improve the postoperative experience of patients.

### 4.2 Risk factor of PCH

Our study divided the PCH of MVD into two dimensions for analysis: severity and duration. Age (55.8±8.3VS50.1±8.2, OR=0.930, 95%CI: 0.870-0.993,P=0.029) and gas in the prepontine cistern (25.0%VS65.5%, OR=3.994, 95% CI: 1.405-11.355,P=0.009) are independent risk factors for the severity of PCH. At the same time, age (54.3±7.8VS47.8±8.7, OR=0.898, 95%CI=0.821-0.983,P=0.02), SAS score(31.4±5.3VS28.9±5.4, OR=1.193,95%CI=1.064-1.338,P=0.002), gas in the prepontine cistern (35.6%VS82.1%, OR=5.513,95%CI=1.423-21.359, P=0.013) and temperature>38.0°C(13.6%VS50.0%, OR=5.326,95%CI=1.305-21.744,P=0.020) are predictive of the duration of PCH independent risk factors.

#### 4.2.1 Age

Age was an independent risk factor for the severity and duration of PCH in our study. This finding was consistent with recent evidence on the risk factors for PCH(Rimmer, Scott et al. 2021). Animal experiments have shown that the number and volume of sensory nerve cells will gradually reach their peaks with age, and then gradually decrease and shrink with age(Cecchini, Cuppini et al. 1995, Gagliese and Melzack 2000). Similar conclusions have been demonstrated in human experiments. Some studies have observed that patients’ tolerance to pain increases with age(Woodrow, Friedman et al. 1972, Devor 1991, Baron 2000, Gibson and Farrell 2004). Our study showed that the average age of the mild group was significantly higher than that of the severe group (55.8±8.3VS50.1±8.2, P=0.003), while the average age of patients in the short-term PCH group was significantly higher than that of the long-term PCH group (54.3±7.8VS47. 8±8.7, P=0.001). Therefore, we consider that for patients younger than 50 years old, we should pay more attention to the patient’s headaches after the operation, and timely and reliable analgesic measures should be given, if necessary, to reduce the degree of pain and the duration of pain after the operation.

#### 4.2.2 SAS scores

Several studies have indicated that anxiety is a risk factor for PCH(De Benedittis, Lorenzetti et al. 1996, Rocha-Filho, Gherpelli et al. 2007, Rocha-Filho 2015, Rimmer, Scott et al. 2021). Consistent with this, the present study found that patients with high SAS scores had a significantly higher duration of the presence of postoperative pain than those with low SAS scores. It is worth noting that the SAS scores of the patients in this study were all less than 50, so none of the patients could be assessed with an anxiety disorder before surgery. Although patients cannot be diagnosed with an anxiety disorder before surgery, we believed it can still reflect that the psychological conditions of patients in the long-term PCH group are different from those in the short-term PCH group. Through the analysis of the various sub-items of SAS, we found that the options with higher patient scores mainly focus on sleep disorders. Patients who experience sleep disturbance may develop negative emotions such as anxiety, loneliness, and irritation(Fadayomi, Ibala et al. 2018). We speculate that perioperative administration of appropriate hypnotics can help shorten postoperative pain duration, which needs further exploration.

#### 4.2.3 Gas in the prepontine cistern area

Previous research showed that the loss of cerebrospinal fluid and cerebrospinal fluid leakage caused by posterior fossa surgery was one of the causes of PCH(Ellis, Speed et al. 1998). The present study believed that intraoperative cerebrospinal fluid loss did not affect the degree and duration of postoperative pain. The total amount of CSF loss in patients was estimated by calculating their 4-hour postoperative intracranial air volume. There was no difference in CSF loss between the mild and severe groups (8.8 ± 6.4vs9.4 ± 7.1, P = 0.656), nor between the short - and long-term PCH groups (8.9 ± 6.5vs9.8 ± 7.5, P = 0.563). This indicates that the lost cerebrospinal fluid will be quickly replenished within 24 hours after surgery.

Interestingly, this study found that it is not the amount of gas but the location of gas that affects postoperative pain. The main pain receptors for headaches are found in the dura mater, periosteum, and scalp tissues, but not brain tissues. However, the main afferent nerves are the trigeminal nerve and the cervical 1-3 nerves(de Gray and Matta 2005). Therefore, we speculate that gas accumulation at the root of the trigeminal nerve rather than the frontal and temporal regions may be a key factor leading to postoperative pain in patients. Through univariate analysis, it was found that the number of patients with gas accumulation in the prepontine cistern in the severe group was significantly higher than that in the mild group. At the same time, regression analysis indicated that gas accumulation in the prepontine cistern was an independent risk factor for pain (OR=3.665, 95% CI: 1.321-10.170). We further analyzed the amount of air accumulation in the prepontine cistern of the two groups of patients and found that the amount of air accumulation in the prepontine cistern in the severe group was significantly higher than that in the mild group (0.11±0.26 VS 0.40±0.61, P=0.003). Regression analysis found that the amount of air in the prepontine cistern is an independent risk factor for the severity of pain(OR=3.91, 95%CI: 1.16-13.16, P=0.028). The trigeminal nerve emerges from the brainstem and is soaked in the prepontine cistern. Therefore, we speculate that the presence of gas in the root of the trigeminal nerve will affect the physical and chemical properties of the root of the trigeminal nerve, and at the same time cause the trigeminal nerve to receive certain mechanical stimulation. The greater the amount of air in the root of the trigeminal nerve, the more severe the harassment of the trigeminal nerve, and therefore the more severe the postoperative pain. Studies have shown that mechanical or chemical stimulation of the trigeminal nerve is one of the key factors of pain after craniotomy(Zhao and Levy 2014, Brazoloto, de Siqueira et al. 2017). However, no research has been found to investigate whether the gas at the root of the trigeminal nerve can cause headaches. Therefore, our speculation needs to be further verified.

### 4.3 Treament for PCH

PCH not only causes discomfort to the patient but is also potentially destructive. It also delays recovery and increases the length of hospital stay. Studies have shown that acute PCH can lead to increased blood pressure, emotional agitation, and vomiting(Hughey, Lesniak et al. 2010). Not only will it cover up symptoms such as increased intracranial pressure and cerebral hemorrhage, but it will also seriously affect the surgeon’s judgment of the patient’s postoperative condition. At the same time, it may increase the risk of postoperative intracranial hematoma(Basali, Mascha et al. 2000, Jian, Li et al. 2014), thereby prolonging the hospital stay(Leslie, Troedel et al. 2003). Acute PCH may induce persistent PCH(Flexman, Ng et al. 2010), and early analgesic measures can help reduce the occurrence of persistent PCH(da Cruz, Moffat et al. 2000). Therefore, it is meaningful to take effective treatments for PCH of MVD.

For PCH, there is currently no unified treatment standard. We review the relevant literature and summarize pain management into three aspects: psychological intervention, drug intervention, and improved surgical methods. Psychological intervention is aimed at young people with high SAS scores, informing in advance of possible expected pain, PCH treatment options, and side effects of analgesics. Studies have shown that psychological intervention can reduce the pain experience of patients (Kastanias, Denny et al. 2009, Kol, Alpar et al. 2014, van Dijk, van Wijck et al. 2015). A drug intervention can be comprehensively applied before, during, and after MVD. This study showed that the use of sodium valproate during the perioperative period could reduce the duration of PCH. However, its mechanism needs to be further explored. Preoperative use of gabapentin(Türe, Sayin et al. 2009), intraoperative use of dexmedetomidine, and postoperative use of non-steroidal anti-inflammatory drugs (NSAIDs)(Galvin, Levy et al. 2019), codeine(Watson 2011), and sumatriptan(Venkatraghavan, Li et al. 2016) can effectively reduce PCH. Intraoperative scalp block and incision infiltration anesthesia can effectively reduce headaches within a few hours after surgery, and may reduce the occurrence of chronic PCH(Jia, Zhao et al. 2019). Improved surgical methods are mainly reflected in the design of different skin incisions. Some studies believe that the PCH of MVD is caused by the over-dissection of local muscles caused by the straight incision and the damage to the minor occipital nerve and the greater auricular nerve(Schessel, Rowed et al. 1993, Schaller and Baumann 2003). Studies have proposed that the modified incisions of C-(Chibbaro, Cebula et al. 2018) and S-shape(Tomasello, Esposito et al. 2016) can not only avoid damage to the minor occipital nerve but also avoid local skin and muscle damage caused by excessive use of mastoid spreaders, thereby reducing the postoperative incidence of headaches.

### 4.4 Limitation

There are several limitations to this study. This is a single-center retrospective evaluation at an institution. Since the sample size was determined by the number of patients in the inclusion period, the sample size was small. There is a certain error in the time point of the patient’s head CT recheck, so there is an error in the statistics of intracranial air volume. The presence of air in the prepontine cistern can cause PCH is an important finding of this study. However, it has not been analyzed for the accumulation of air in the frontal and temporal regions. We will further improve related research and explore the mechanism of PCH caused by gas accumulating in the prepontine cistern area.

## 5. Conclusion

PCH is a common complication after MVD surgery among patients with HFS, occurring in more than half of our patients (63.2%). At the same time, this is a self-limiting complication that will gradually disappear within 7 days after the operation. Young age, temperature > 38°C after MVD within 24h, and gas around the prepontine cistern are associated with developing the severity and duration of PCH after MVD procedures. Patients with these risk factors should be closely monitored and should be given appropriate analgesic measures during the perioperative period.

## Data Availability

All relevant data are within the manuscript and its Supporting Information files.

## 6. Disclosures and Acknowledgments

Nothing to disclose.

This research did not receive any specific grant from funding agencies in the public, commercial, or not-for-profit sectors.

